# An Atomic Approach to the Design and Implementation of a Research Data Warehouse

**DOI:** 10.1101/2021.05.05.21256679

**Authors:** Shyam Visweswaran, Brian McLay, Nickie Cappella, Michele Morris, John T. Milnes, Steven E. Reis, Jonathan C. Silverstein, Michael J. Becich

## Abstract

**Objective:** As a long-standing Clinical and Translational Science Awards (CTSA) Program hub, the University of Pittsburgh and the University of Pittsburgh Medical Center (UPMC) developed and implemented a modern research data warehouse (RDW) to efficiently provision electronic patient data for clinical and translational research.

**Methods:** Because UPMC is one of the largest health care systems in the US with multiple vendors’ electronic health record (EHR) systems, we designed and implemented an RDW named Neptune to serve the specific needs of our CTSA. Neptune uses an atomic design where data is stored at a high level of granularity as represented in source systems. Neptune contains robust patient identity management tailored for research; integrates patient data from multiple sources, including EHRs, health plans, and research studies; and includes knowledge for mapping to standard terminologies. Neptune enables efficient provisioning of data to large analytics-oriented data models and to individual investigators.

**Results:** Neptune contains data for more than 5 million patients longitudinally organized as HIPAA Limited Data with dates and includes structured EHR data, clinical documents, health insurance claims, and research data. Neptune is used as a source for patient data for hundreds of IRB-approved research projects by local investigators and for national projects such as the Accrual to Clinical Trials (ACT) network, the All of Us Research Program, and the National Patient-Centered Clinical Research Network.

**Discussion:** The design of Neptune was heavily influenced by the large size of UPMC, the varied data sources, and the rich partnership between the University and the healthcare system. It features several desiderata of an RDW, including robust protected health information management, an extensible information storage model, and binding to standard terminologies at the time of data delivery. It also includes several unique aspects, including the physical warehouse straddling the University of Pittsburgh and UPMC networks and management under a HIPAA Business Associates Agreement.

**Conclusion:** We describe the design and implementation of an RDW at a large academic health care system that uses a distinctive atomic design where data is stored at a high level of granularity.

## INTRODUCTION

The passage of the Health Information Technology for Economic and Clinical Health (HITECH) Act by the United States federal government led to the widespread adoption of electronic health record (EHR) systems that capture patient data at an ever-increasing pace (1). The availability of large amounts of EHR data provides new opportunities for their secondary use to support clinical and translational science. Further, EHR data in combination with other patient data from research studies, patient-reported outcomes, mobile health, and social media are progressively becoming important in biomedical research.

Data warehouses containing EHR data exist in large healthcare systems for a variety of operational, reporting, quality improvement, and financial purposes (2). However, such warehouses often do not support research effectively due to the heterogeneity of EHR data, regulatory complexity such as the requirement for de-identification (3), and the need for research-project-specific data management. A common approach to efficient and large-scale reuse of EHR data for research is a dedicated research patient data repository or research data warehouse (RDW) that integrates and harmonizes EHR data and is architected, implemented, and operated by personnel with informatics expertise. Funded by the National Center for Advancing Translational Sciences (NCATS), the Clinical and Translational Science Awards (CTSA) Program hubs have developed RDWs for efficient and widespread use of EHR and other data for research; with 94% of all hubs providing such services (4).

Dedicated RDWs have enabled a wide range of research efforts such as clinical trial recruitment, large-scale characterization of treatment pathways (5), generation of real-world evidence for clinical decision making (6), pharmacovigilance (7), rapid cohort identification (8), and phenome-wide association studies (9). Furthermore, harmonized data in RDWs unlock future opportunities for large scale application of machine learning for biomedical discovery (10) and clinical decision support that can support order entry (11), smart prioritization of data in EHR systems (12), anomaly detection (13), and precision medicine (14).

RDWs have evolved along two broad pathways (15). Several large academic health centers have developed single institutional RDWs that are architected specifically based on local EHR systems and needs. Examples of single-institution RDWs are those at Northwestern University (16, 17), Duke University Health System (18), Stanford University (19, 20), and Vanderbilt University (21). Other institutions have implemented RDWs based upon analytics-oriented data models designed for multi-institutional consortia and data networks. Examples of such data models include the Informatics for Integrating Biology and the Bedside (i2b2) (22), the Observational Medical Outcomes Partnership (OMOP) Common Data Model (23), and the National Patient-Centered Clinical Research Network (PCORnet) Common Data Model (24).

In this paper, we describe the design and implementation of a single institutional RDW, called Neptune, at the University of Pittsburgh (Pitt). Neptune is architected to ingest patient data from a multitude of sources, to store data at the level of granularity that exists in the sources, and from which data is subsequently transformed into analytics-oriented data models and research data sets. Beyond patient data, knowledge for mapping to standard terminologies and definitions for standardizing clinical concepts are also stored in Neptune. We provide a brief description of the large health system associated with Pitt, details of the architecture of Neptune, and some of the distinctive aspects of Neptune related to the technical infrastructure.

## METHODS

### Setting and history

The University of Pittsburgh Medical Center (UPMC) is one of the largest health care systems in the United States. UPMC serves western, central, and western Pennsylvania and parts of Ohio, West Virginia, and New York, and comprises 40 hospitals with 8,400 licensed beds, more than 700 doctors’ offices and outpatient facilities, and 23 nursing homes. Annually, UPMC has 388K inpatient admissions, 1.1M emergency room visits, 5.5M outpatient visits, and 260K surgical procedures. The University of Pittsburgh School of Medicine (UPSOM), located in the city of Pittsburgh, is the medical college and the clinical research facility that, together with UPMC, comprises a top 5 NIH-supported leading academic medical center. UPSOM supports an academic staff of nearly 2,500 physicians and educators and trains approximately 600 medical students and 1,900 medical residents and clinical fellows yearly.

UPMC has evolved as a merger of previously independent hospitals and practices, and its clinical information systems reflect this heritage, including a range of legacy and modern systems. UPMC has deployed several EHR systems from different vendors. In most outpatient facilities, UPMC uses the EpicCare system (Epic, Verona, Wis.), while in the inpatient and emergency settings, UPMC has deployed the Cerner system. The UPMC Children’s Hospital of Pittsburgh has an independent installation of the Cerner Millennium system. Additional EHR and ancillary systems are used in various specialty settings such as inpatient psychiatry, the cancer center, the perioperative setting, and radiological imaging. UPMC has created multiple interfaces among the clinical information systems to enable clinical workflows that require data from multiple systems; however, this has led to the replication of patient data across these systems.

As early as 1982, Pitt and UPMC developed a clinical data warehouse called the Medical ARchival Retrieval System (MARS) that integrated data from EHR systems and administrative claims systems (25). MARS was developed as a file-based database system that archived both structured and document patient data in text files. As UPMC grew with the acquisition of hospitals and their clinical information systems, data integration was achieved by sending data in Health Level Seven (HL7) format through a message router to MARS.

Since MARS was implemented two decades ago, UPMC has grown substantially and has deployed several modern EHR systems. The need for a modern dedicated RDW emerged over the past several years. As a long-standing CTSA hub, Pitt needed a modern RDW and robust informatics services for efficient and effective support of investigators at Pitt and UPMC.

### Organization

The Biomedical Informatics Core (BIC) of the University of Pittsburgh Clinical and Translational Science Institute, the Center for Clinical Research Informatics (CCRI) (26), and the Research Informatics Office (RIO) (27) lead the development of Neptune and provisioning of patient data for research. BIC, CCRI, and RIO are each housed in the Department of Biomedical Informatics and are each led by informatics faculty. On behalf of UPMC, three informatics faculty members oversee long-term planning, implementation of new features, and maintenance of Neptune and its downstream analytics-oriented data marts for national data-sharing efforts.

The team that supports Neptune and the data marts perform a range of functions. The *technical group* identifies use cases, reviews and selects technologies, implements extract-transform-load processes based on data standards and terminologies established by a data harmonization group, and develops processes for aligning and integrating research data with the EHR data. The *data harmonization group* establishes data standards for different types of EHR and research data, determines which standard terminologies to use, and maintains and updates mappings of local terms to terminologies. The *data quality group* establishes statistics, uncovers data anomalies by periodically measuring these statistics in the data, and returns discoveries to the technical group for changes in the data pipelines. The *user support group* communicates with local users, provides training and support, and solicits feedback from the user community.

### Architecture of Neptune

The Neptune RDW consists of three main layers: 1) an *identity management layer* for managing personally identifiable information, 2) a *data layer* that contains EHR and other patient data, both identified and as limited data with preserved timestamps and zip codes, and 3) a *semantic layer* that consists of business logic such as mappings between local terms and standard terminologies (see Figure 1). Neptune uses a normalized atomic design. Normalized data implies that one fact or piece of information is stored in one place in the warehouse to minimize redundancy. An atomic data warehouse (28) contains data at a high level of granularity and is obtained from the source systems with minimal filtering or summarization.

**Figure 1.**
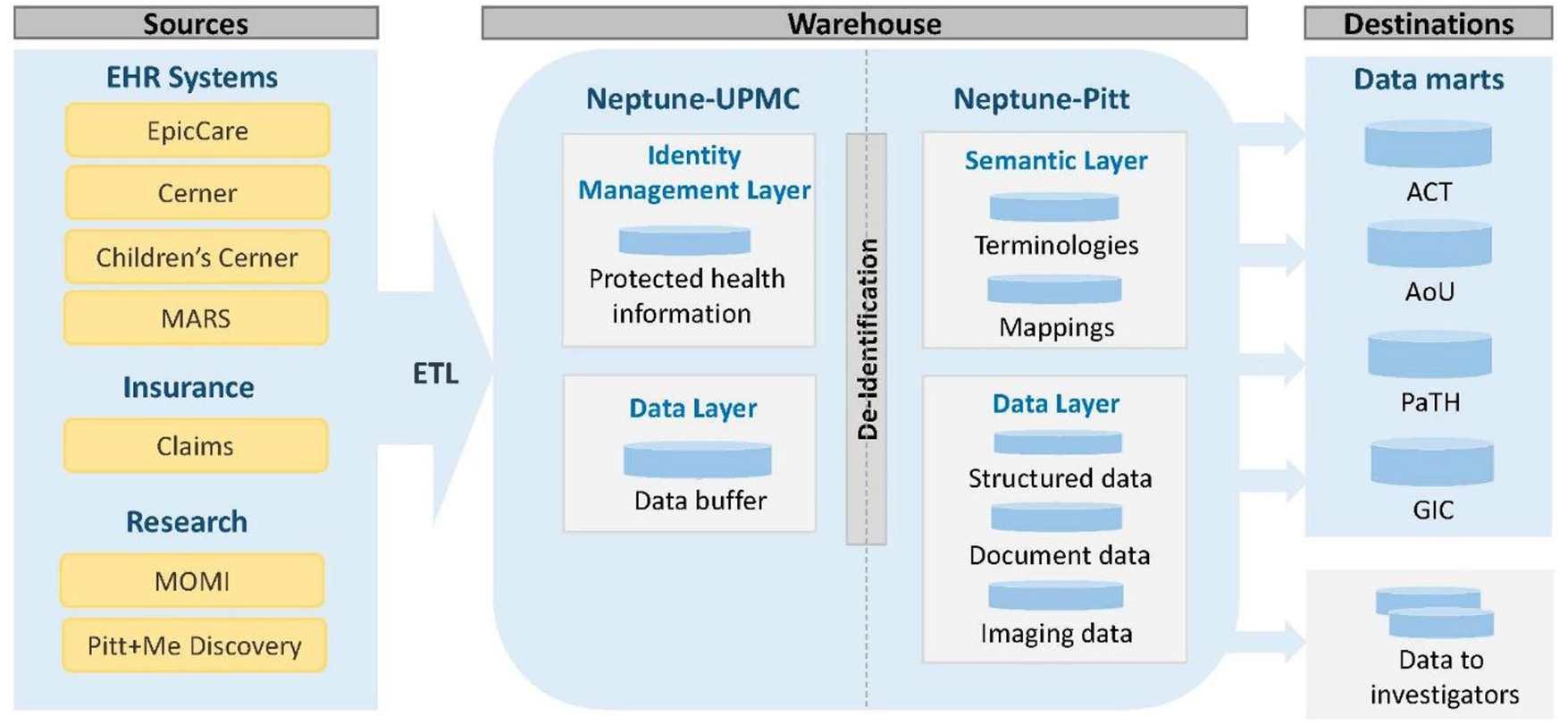
The architecture of Neptune with sources and destinations. The identity management layer resides at UPMC, the semantic layer resides at Pitt, and the data layer resides mostly at Pitt.

The RDW is implemented using the Oracle database management system. The warehouse physically straddles the UPMC and the Pitt networks. For example, the identity management layer of Neptune resides within the UPMC network, and the semantic layer and most of the data layer resides in the Pitt network (see Figure 1). Though the warehouse is split across two distinct networks, members of the technical group can seamlessly view tables from both components of Neptune and run processes across all layers of Neptune.

#### Identity management layer

The identity management layer resides in the UPMC network and contains personally identifiable information of all patients. A key function in Neptune is to assign and maintain a unique research enterprise identifier to each patient. This research enterprise identifier is distinct from patient identifiers that are used in the healthcare system, including clinical enterprise identifiers, medical record numbers, and other healthcare identifiers. The research enterprise identifier is linked to healthcare system patient identifiers and is also linked to participant identifiers of research data sets that are integrated into Neptune. This three-layer identity management exceeds best practices for HIPAA Honest Brokerage and helps ensure participant identifiers cannot be shared across projects.

Identity management and linking of patient identifiers is performed in a staging area. During monthly ingestion of data from clinical systems, new patients in the health system are identified and assigned new research enterprise identifiers. For existing patients who may have been assigned new clinical identifiers, the new identifiers are linked to the existing research enterprise identifier. Any merges of clinical enterprise identifiers and medical record numbers are also processed. Identity management enables patient data from any data domain and linked to any patient identifier to be accurately linked to the enterprise research identifier. Neptune’s identity management achieves a key goal of Neptune: to create a comprehensive longitudinal record for each patient by integrating clinical and non-clinical data from multiple sources.

#### Data layer

The data layer resides mostly at Pitt and contains both structured and text EHR data as well as other types of data such as imaging data (see Figure 2). The data layer stores atomic patient data; that is, the data is at the level of granularity in the source system with minimal transformation. The structured data include core data domains such as demographics, visits, diagnoses, procedures, laboratory test results, medication orders, other orders (for laboratory tests, clinical imaging, procedures, etc.), and medication dispenses. Additional data domains include allergies, vaccine administrations, and metadata of clinical documents. The text data consists of de-identified content for all document types such as history and physical, progress, consultation, procedure, and discharge notes; radiology and pathology reports; electrocardiogram and electroencephalogram reports; and many more. In every domain, for each data item, the source system from which it was extracted is recorded to maintain data provenance.

**Figure 2.**
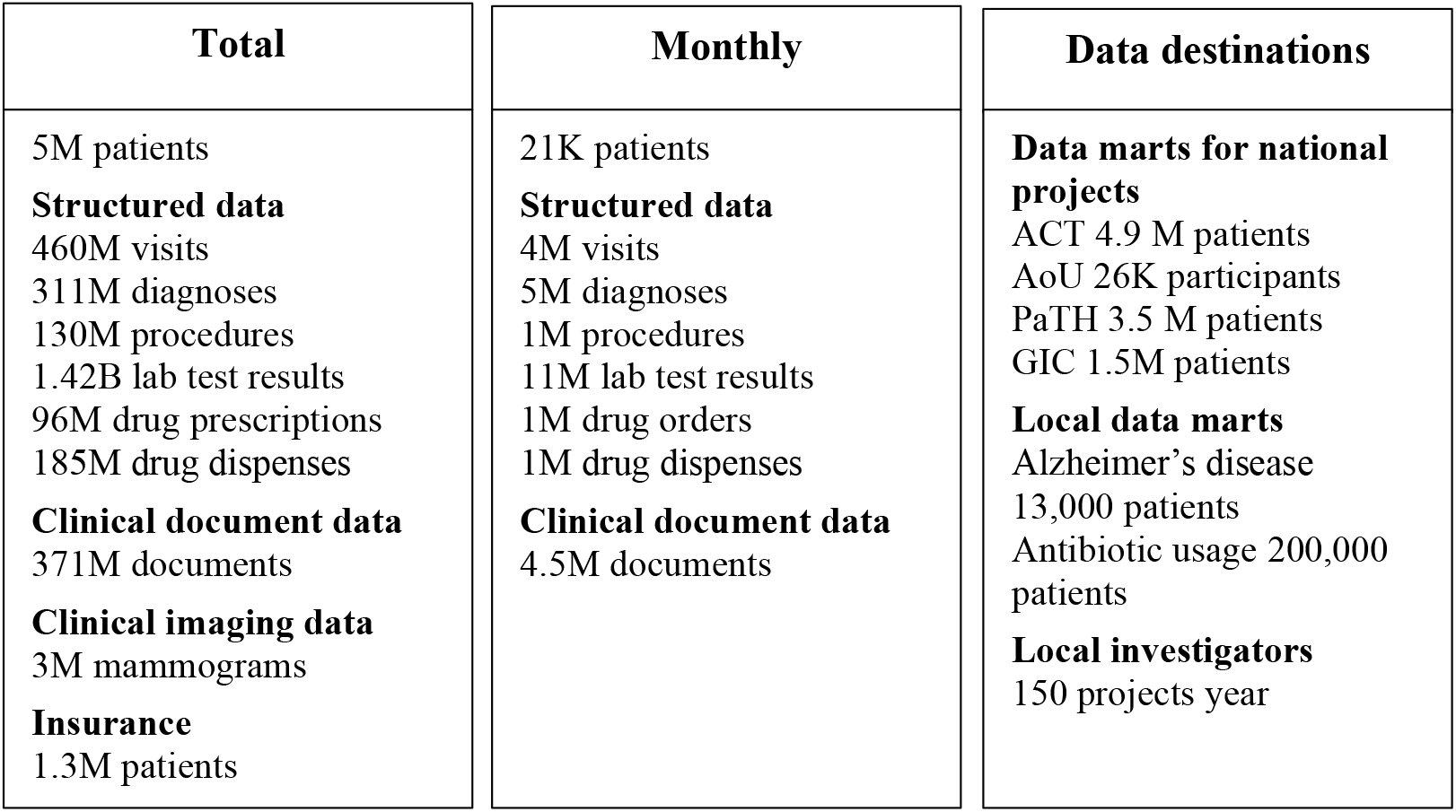
Total volume of data in Neptune, monthly data inflows and data volumes in destinations served by Neptune.

At the time of extraction of data from source systems, the extracts are staged in the data layer at UPMC, where de-identification is performed before data is moved to the data layer at Pitt. A copy of the latest extract of data for all domains is maintained in the staging data layer. For most domains, data is extracted and processed monthly. De-identification consists of removing all HIPAA specified personally identifiable information with the exception of dates and zip codes to create a Limited Data Set, and the data are linked to the patient only through the research enterprise identifier. De-identification is straightforward for structured data domains where database columns containing personally identifiable information are removed. De-identification of clinical documents is done using NLM Scrubber (29) that has been adapted for our use.

#### Semantic layer

The semantic layer resides at Pitt and contains the knowledge and logic to harmonize data that may be represented in a heterogeneous fashion across different hospitals and source systems. For example, for the laboratory test of hematocrit, each hospital at UPMC uses a different local code, and the semantic layer contains a list of all local hematocrit codes that are mapped to the relevant LOINC code for standardization. Mappings are obtained from several sources. One source is reference data obtained from source systems like EpicCare that contains mappings between local terms and standard terminologies that are created and maintained by the clinical enterprise. However, the clinical enterprise does not necessarily create mappings to legacy data or mappings to standard terminologies that are not mandated by federal regulations. The data harmonization group creates and updates mappings for legacy data and mappings that are useful in research. In addition, we import a comprehensive set of medical terminologies that are contained in the Unified Medical Language System (UMLS) (30) and maintain all versions of the terminologies going back to 2004 UMLS releases. We harmonize diagnoses to ICD-9 and ICD-10, procedures to ICD-9, ICD-10, CPT-4 and HCPCS, medications to RxNorm and NDC, and laboratory tests to LOINC. We also use the UMLS to validate the source system data that are increasingly coded with standard terminologies. In addition to mappings, the semantic layer contains value sets that have been collected from several sources such as the NIH’s Value Set Authority Center (31) and value sets that have been defined by national patient data research networks.

The mappings and value sets of the semantic layer are leveraged at the time data is delivered from Neptune to downstream data marts and to individual projects. Standard terminology codes and values are applied at that time to produce standardized data; this late binding approach provides efficient and timely standardization of data to constantly changing terminologies.

### Extract, transform, and load processes

Most data domains in Neptune are updated at the beginning of each month. A series of extract, transform, load (ETL) database operations are implemented using the Pentaho Data Integrator (PDI) and run overnight to extract a month of data from the source systems. More than 70 workflows process the monthly incremental data updates. The PDI programs perform extraction and loading of warehouse tables, including data validation checks, error handling, auditing, and control processing. Linux scripts are used to call PDI programs. The data in the source systems are transactional, and some transactions may take several weeks to be finalized. Thus, we update data in Neptune with a lag period of at least one calendar month so that once extracted and loaded, data in Neptune do not have to be updated nor reconciled with newer data.

### Extension of Neptune for COVID-19

The emergence of coronavirus disease 2019 (COVID-19) necessitated a more frequent update of COVID-19 related data in Neptune to support surveillance and research needs. Since UPMC is a large health system with millions of active patients and billions of transactions, rather than changing the monthly ETL processes, we developed a new parallel ETL process that updates data on COVID-19 twice a week. For efficiency, the new process extracts data only between the latest monthly Neptune update and the current day; thus, the data lags the source systems at most by four days. This COVID-19 component of Neptune serves national projects that added more frequent data requirements for COVID-19 serving the COVID-19 needs of local investigators.

### Regulation of Neptune

Neptune was initially developed under an institutional review board (IRB) protocol, but the regulatory framework was later changed to a HIPAA Business Associates Agreement (BAA). Regulation under an IRB protocol was limiting since the addition of new data sources, and technical changes need repeated changes to the protocol. The BAA is overseen by the Chief Research Informatics Officer at Pitt and the Chief Medical Information Officer at UPMC. This arrangement, allowing Neptune to function as an operational research system of UPMC under the BAA, enabled rapid expansion and development of new functionality in Neptune.

## RESULTS

Designed as a RDW that integrates patient data from varied sources, Neptune contains EHR data (structured, document and imaging), insurance data, and research data. EHR data in Neptune goes back to 2004 when UPMC completed implementation of electronic clinical information systems. Every month, a large volume of EHR data is added to Neptune that includes data from existing patients and approximately 21,000 new patients (see Figure 2). Neptune also receives health insurance claims data from the UPMC Health Plan and from large institutional research projects like the Magee Obstetric Maternal & Infant (MOMI) Database and Biobank (>300 perinatal variables from mother and infant, ∼200K deliveries since 1995) (32, 33), and genomic data from the Pitt+Me Discovery Biobank.

Neptune provides data to data marts for several national projects that include the NCATS-funded Accrual to Clinical Trials (ACT) network (8, 34), which is based on i2b2 (22) and SHRINE (35); the NIH-funded All of Us Research Program which is based on the OMOP data model (36, 37); the PCORI-funded PCORnet, which is based on PCORnet’s Common Data Model (CDM) and

PopMedNet (38, 39); and the NCATS-funded Genomic Information Commons (GIC) which is based on i2b2/TranSMART (40, 41). Neptune also provides data to data marts for local projects such as an Alzheimer’s disease project and an antibiotic usage project. Typically, data is automatically updated in both the national and local data marts following the monthly data updates in Neptune (see Figure 2).

In addition, Neptune serves as a source of EHR and other patient data for local research in the institution. The RIO provisions data to hundreds of individual research projects per year. Finally, RIO responds to approximately one thousand requests per year, including preparatory to research requests and letters of support for research grants.

## DISCUSSION

We described the design and implementation of Neptune, a new RDW, at Pitt and UPMC. Neptune is designed to integrate data from several EHR systems with replicated patient records as well as non-EHR data, support both identified and de-identified data needs, and service efficiently commonly used analytics-oriented data models and data needs of individual investigators. The rich partnership between Pitt and UPMC supported the rapid technical development and implementation of Neptune. This warehouse is an increasingly rich repository of EHR and other patient data and is progressively benefitting the dynamic research environment at Pitt and UPMC.

### Distinctive features of Neptune

This section describes distinctive aspects of Neptune ‘s technical infrastructure. Desiderata for the successful implementation and operation of an RDW has been described by Huser and Cimino (42). These include a single patient identifier, protected health information (PHI) management, an extensible information storage model, semantic integration with standard terminologies, metadata and documentation, and documentation of historical evolution of data sources. Several of the features of Neptune described below align with these desiderata.

#### Atomic data warehouse

Neptune is architected as a canonical model for ingestion and storage of patient data derived from multiple sources and from which data is subsequently transformed into analytics-oriented data models. The canonical model in Neptune uses a normalized atomic design. Normalization is a key database principle that enables efficient correction of data errors and optimization of storage space. The atomic design enables rapid ingestion of data in bulk, tracking of data provenance, isolation, separate processing of changing data, and provides a single place for data cleaning and transformation rather than duplicating these processes for each data source. The advantage of an atomic warehouse is it can both provide answers to queries at a very detailed level and summarize data rapidly that may be needed for analytics-oriented data models. Neptune enables us to avoid converting data from one data model to another, e.g., from OMOP to PCORnet’s CDM or vice-versa, which is typically more complex to implement than an ETL process from Neptune to an individual data model. Further, due to information loss, it is not possible to inter-convert between data models with complete fidelity. The atomic design also enables the stepwise addition of new data domains without the need to redesign nor implement a comprehensive set of all possible data domains that will eventually be needed.

#### Single patient identifier and management of PHI

A key feature in Neptune is the management of patient identifiers such that all data related to a patient originating from different EHR systems, health insurance, and research studies are linked to a single enterprise research identifier. While UPMC maintains a single enterprise clinical patient identifier that links all clinical identifiers of a patient, the enterprise clinical patient identifier was not usable in the RDW for several reasons. We needed an enterprise identifier that is not PHI and can be linked to both clinical patient identifiers and patient identifiers used in research studies. In Neptune, the identity management layer is used to integrate clinical and non-clinical identifiers and assign a unique enterprise research identifier to each patient. This layer resides on the health system side since it contains PHI; thus, the architecture of Neptune helps ensure that PHI does not leave the confines of the health system network.

#### Privacy and study patient identifiers

The enterprise research patient identifier is restricted for use within Neptune and is not used to identify patients when data is delivered to data marts and for research projects. Unique study patient identifiers are created and assigned for patients in each data mart and research data set that are derived from Neptune. A function is used to systematically transform the enterprise research patient identifier to a study patient identifier for each data set, and function details associated with each data set are archived in Neptune. The function allows warehouse personnel to link study identifiers to enterprise research patient identifiers for future updates to study data, but investigators cannot link data by patient across different data sets that were provisioned under different IRB protocols that may have common patients.

#### Extensible information storage model

An important consideration for Neptune was rapid implementation, starting with key data domains so that the warehouse would be functional within months rather than years of development. The key data domains were identified based on the clinical domains required to service the national data marts; as such, Neptune initially contained only structured EHR data in the domains of demographics, diagnoses, procedures, laboratory test results, and medications. This enabled implementation in under six months. The addition of a new domain includes a selection of sources, identification of de-duplication strategies if necessary, aligning patient identifiers, a bulk backload of the data going back to 2004, and implementation of a monthly ETL process. The extensible information storage model implemented in Neptune has enabled the stepwise addition of new data domains without the need to rearchitect existing data domains.

#### De-replication of data from multiple sources

Multiple EHR systems and an archival system are in use in UPMC. Assembling a longitudinal health record from these multiple sources is another key requirement for Neptune. In addition to the multitude of patient identifiers, another challenge associated with the use of multiple EHR systems is the replication of patient data across systems. We achieved de-replication in several ways. One approach is the selective extraction of data from a single source if a particular domain is systematically replicated across the EHR systems. For example, since 2015, in UPMC, laboratory test results from all care settings are available in EpicCare; thus, laboratory test results after 2015 are extracted only from EpicCare. Another approach compares timestamps and metadata of suspected replicated data to identify replication. For example, laboratory test results before 2015 were obtained from several sources, and replications were identified and systematically eliminated.

#### Binding at query

Binding is the process of mapping data to standard terminologies (e.g., translation of a local code for a laboratory test to the appropriate LOINC code) and application of definitions (e.g., application of a standard definition of an outpatient visit and calculation of the length of stay). Binding standardizes the data and makes it usable for research. In some warehouse designs and analytics-oriented models, mappings and definitions are applied early during data ingestion; such early binding has the disadvantage that changes to the mappings and definitions will need data to be corrected and updated continually. Since Neptune uses binding at query time, changes in mappings and definitions affect data only at the time data is delivered from Neptune, and new data sources are rapidly integrated into Neptune without making decisions about mappings upfront.

### Limitations

Neptune has several limitations. One limitation is that the data in the warehouse lags the source systems by a month. While this delay is acceptable for most research that uses retrospective data, it limits research in clinical decision support and biosurveillance applications that typically require current or near current EHR data. But, as mentioned previously for COVID-19 data, extending the capabilities to support requirements of more frequent data updates is possible with additional development. Another limitation is that there is no efficient mechanism to query clinical document data, while structured data can be queried by the warehouse personnel by directly querying Neptune or via the ACT i2b2 data mart. We have separately implemented Elasticsearch technology for efficient query and analysis of text documents.

## CONCLUSION

The Neptune RDW implemented at Pitt is increasingly enabling extensive reuse of patient data for a wide range and high volume of clinical and translational research. Neptune is designed as a normalized atomic warehouse. The atomic design enabled the warehouse to be built “better, faster, cheaper” because there is no need to extensively model or standardize the data. Neptune integrates patient data from multiple EHR systems as well as from other sources, maintains a robust patient identity management system for research, and enables efficient delivery of data to both large data marts based on analytics-oriented data models and to individual investigators. Creating a dedicated RDW at Pitt has enabled us to better serve the investigators at Pitt, participate in regional and national data networks, and advance informatics research.

## Data Availability

The data in the patient data warehouse described in the article cannot be shared publicly due to the privacy of individuals.

## Acknowledgments

The research reported in this article was supported by awards from the National Center for Advancing Translational Sciences of the National Institutes of Health (NIH) under award numbers UL1 TR001857, UL1 TR001857-01S1 and U01 TR002623, the Office of the Director of the NIH under award number OT2 OD026554, the National Library of Medicine of the NIH under award number R01 LM012095, and the PCORnet PaTH network RI-CRN-2020-006. The content is solely the responsibility of the authors and does not necessarily represent the official views of the NIH.

